# Parent’s Views on Access to Dental Care and the Interim Canadian Dental Benefit

**DOI:** 10.1101/2024.05.10.24307141

**Authors:** Anil Menon, Vivianne Cruz de Jesus, Jorma I. Virtanen, Robert J. Schroth

## Abstract

**Introduction:** This study investigated parents’ perspectives on access to oral health care and the Interim Canada Dental Benefit (CDB). In the context of Canada’s national health insurance, which historically excluded dental care, the introduction of the Interim CDB in October 2022 represented a paradigm shift towards enhancing dental care accessibility for children under 12 years of age from lower-income families.

**Methods:** This study analyzed aggregate and de-identified data from the comprehensive online survey conducted by The Strategic Counsel for Health Canada, involving 2,203 parents from across Canada. The survey was administered in March of 2023. Paired/overlap t-test for means and paired/overlap z-test for percentages were performed, with statistical significance at *p* ≤ 0.05.

**Results:** The majority of participants expressed concerns regarding the costs (90.9%) and accessibility (80.9%) of dental care, indicated that regular dental visits for children is important (97.2%), and would take their children more frequently to dental appointments if had extra money (79.9%). Some of the barriers preventing regular dental visits for children included costs of service and transportation and lack of insurance. The majority of parents showed support for the Interim CDB (87%), with the greatest support coming from the provinces of Manitoba and Saskatchewan (90.4%).

**Conclusion:** This research underscores the imperative for ongoing evaluation and policy refinement to ensure the CDCP effectively addresses the nuanced needs of Canadian families, fostering a more inclusive and accessible dental care system. Parents’ concerns regarding dental care and their support for the Interim CDB signal a clear mandate for improving program outreach and accessibility through the Canadian Dental Care Plan (CDCP).

**Knowledge Transfer Statement:** Findings from this study highlight the significant concern among parents regarding dental care affordability in Canada, reinforcing the necessity of programs like the Interim Canada Dental Benefit (CDB) and Canadian Dental Care Plan. High rates of support suggest a positive public reception of the CDB, which is crucial for policy intervention’s success. Concern about accessing dental services, despite the availability of the insurance, indicates ongoing barriers to dental care, suggesting areas for future policy refinements.

## Introduction

Canada’s national health insurance (Medicare) did not historically include dental care, given the country’s history of privileging equal access to health care. The limited public funding in Canada covers the cost of dental care for certain groups of people. The federal government funds dental care services to specific groups, including state-recognized Indigenous populations (registered First Nations and Inuit) and the country’s Armed Forces (Farmer et al. 2022).

Meanwhile, provincial governments finance dental care for low-income children, social welfare recipients, individuals with disabilities, and those with craniofacial disorders (Canadian Academy of Health Sciences 2014). Moreover, municipalities often share the cost of providing care for low-income children and social welfare recipients with the provinces. In addition, low-income seniors receive independent care from municipalities. Despite these government investments, access to oral health care in Canada continues to be a significant challenge (Canadian Academy of Health Sciences 2014; Schroth et al. 2024). As Canada progresses in its efforts to enhance oral health policies, international perspectives are helpful. Free dental care for children is offered in some countries, such as the United Kingdom and the Nordic countries.

Australia’s public dental program provides free or low-cost services to eligible adults and children, with a focus on those with healthcare cards or pensions. Unfortunately, similar to Canada, access to services can be limited due to availability and long wait times, which underscores a common challenge faced by public dental health systems across the globe.

Access to care is a complex issue that includes affordability, availability, accessibility, acceptability, awareness, and accommodation (Saurman 2016). The introduction of the Interim Canada Dental Benefit (CDB) and, more recently, the Canadian Dental Care Plan (CDCP) represent a significant stride towards achieving equitable dental care in Canada. These initiatives are designed to break down financial barriers (i.e. address the affordability dimension), thereby enhancing the accessibility of dental services for lower-income families. They also signify a shift towards a more inclusive healthcare system. The feedback from the community, particularly from parents and children, provides invaluable insights into the effectiveness of these programs, thereby identifying areas for further enhancement.

In October 2022, the Canadian government launched the CDB for children < 12 years of age from families with annual incomes < $90,000 without private dental insurance (Canada Revenue Agency 2022; Department of Finance Canada 2022; Government of Canada 2022; Rabson 2023; Rollason 2023). The “cost of living relief for dental care and rental housing bill” provides financial support up to $650 for each child if the family’s adjusted net income is < $70,000, while $390 is provided if the adjusted net income is between $70,000 and $79,999, and $260 if it is between $80,000 and $89,999 (Canada Revenue Agency 2022; Department of Finance Canada 2022; Government of Canada 2022). The Interim CDB is a precursor to the Canadian Dental Care Plan (CDCP). The Interim CDB will sunset June 30, 2024, while the CDCP is scheduled to commence by mid-2024, providing coverage for uninsured Canadians with annual family income < $90,000 (Nadeau 2023).

To receive the Interim CDB, families must have a child or children < 12 years of age at the time of application, no private dental insurance, and an adjusted family net income of < $90,000 per year. Additionally, families must have filed a 2022 tax return with the Canada Revenue Agency (CRA) and incurred out-of-pocket dental care expenses for their child between October 1, 2022, and June 30, 2024 (Nadeau 2023).

Research shows that multiple factors influence children’s use of dental services. These include their parents’ dental care utilization, level of education and socioeconomic status, income, employment status, access to dental care, dental insurance, behavioural beliefs, perceived power, subjective norms, and parental preventative practices (Badri et al. 2014; Isong et al. 2010; Nagdev et al. 2023). Furthermore, studies have shown that racial and ethnic disparities in dental care utilization are significantly reduced when a child has a parent who uses oral health care and has access to childcare and continuous insurance coverage (Guarnizo-Herreno and Wehby 2012).

The purpose of this study was to investigate the perspectives of parents of children under 12 years old on dental care accessibility challenges and the Interim CDB in anticipation of the CDCP.

## Methods

This cross-sectional study analyzed data arising from the public opinion research online survey undertaken by The Strategic Counsel on behalf of Health Canada’s Oral Health Branch in 2023 (Health Canada 2023). A 15-minute online survey was completed by 2,203 parents of children < 12 years of age with an annual household income < $90,000 Canadian. All participants were over 18 years of age.

Quotas were applied to the base sample (*n* = 2,000) to ensure broad representation from all regions of Canada and a more significant proportion (70%) of Canadian parents with no access to private dental care insurance, as this is one of the requirements for eligibility for the Interim CDB (Health Canada 2023). Two additional ‘sample boosts’ (*n* = 100 each) were carried out to ensure good representation from those residing in remote areas (those located over 350 kilometres from the nearest dental professional and/or without year-round road access) and those who identify as being a member of an ethnic minority community (Table 1). Inclusion of parents from rural and remote areas, as well as ethnic minorities, was purposeful to obtain the experiences of often under-researched demographics, contributing to a more inclusive understanding of dental care accessibility.

**Table 1.** Characteristics of the participants.

|  | **Total**<br>**(A)** | **Atlantic**<br>**(B)** | **Quebec**<br>**(C)** | **Ontario**<br>**(D)** | **Manitoba/**<br>**Saskatchewan**<br>**(E)** | **Alberta**<br>**(F)** | **British**<br>**Columbia/**<br>**North**<br>**(G)** | **URBAN**<br>**(H)** | **RURAL**<br>**(I)** |
| --- | --- | --- | --- | --- | --- | --- | --- | --- | --- |
| **Age of parents** | *n* = 2,201 | *n* = 170 | *n* = 480 | *n* = 829 | *n* = 175 | *n* = 264 | *n* = 283 | *n* = 1,787 | *n* = 392 |
| 18-24 | 51<br>(2.3%)<sup>H</sup> | 5 (2.8%) | 11 (2.2%) | 15 (1.8%) | - | 11<br>(4.3%) | 6 (2.0%) | 34 (1.9%) | 18<br>(4.5%)<sup>A</sup><sub>H</sub> |
| 25-34 | 699<br>(31.8%)<sup>E</sup> | 52 (30.6%) | 163<br>(34.1%)<sup>E</sup> | 259<br>(31.3%)<sup>E</sup> | 39 (22.2%) | 99<br>(37.6%)<sup>AE</sup> | 86<br>(30.5%)<sup>E</sup> | 570<br>(31.9%) | 121<br>(30.9%) |
| 35-44 | 1,032<br>(46.9%) | 81 (47.6%) | 221<br>(46.1%) | 388<br>(46.9%) | 87 (49.5%) | 111<br>(42.1%) | 143<br>(50.7%)<sup>F</sup> | 839<br>(46.9%) | 180<br>(45.8%) |
| 45-54 | 360<br>(16.4%) | 25 (14.8%) | 76 (15.8%) | 139<br>(16.7%) | 38 (21.9%)<sup>AF</sup> | 38<br>(14.6%) | 44 (15.4%) | 299<br>(16.7%) | 61<br>(15.6%) |
| 55-64 | 45 (2.1%)<sup>F</sup> | 7 (4.2%)<sup>F</sup> | 7 (1.4%) | 19 (2.3%)<sup>F</sup> | 6 (3.6%)<sup>F</sup> | - | - | 34 (1.9%) | 12 (3%) |
| 65 or older | 13 (0.6%) | - | - | 8 (1%) | - | - | - | 12 (0.7%) | - |
| **Gender** | *n* = 2,201 | *n* = 170 | *n* = 480 | *n* = 829 | *n* = 175 | *n* = 264 | *n* = 283 | *n* = 1,787 | *n* = 392 |
| Male | 829<br>(37.7%)<sup>IC</sup> | 58 (33.8%) | 150<br>(31.3%) | 327<br>(39.5%)<sup>C</sup> | 77 (44.2%)<sup>BC</sup> | 108<br>(41%)<sup>C</sup> | 109<br>(38.4%)<sup>C</sup> | 716<br>(40%)<sup>AI</sup> | 108<br>(27.6%) |
| Female | 1,367<br>(62.1%)<sup>H</sup> | 112<br>(66.2%)<sup>E</sup> | 330<br>(68.7%)<sup>ADE</sup><sub>FG</sub> | 500<br>(60.4%) | 98 (55.8%) | 154<br>(58.3%) | 172<br>(60.9%) | 1,068<br>(59.7%) | 283<br>(72.1%)<sup>AH</sup> |
| Other or prefer not to answer | 5 (0.2%) | - | - | - | - | - | - | - | - |
| **Education** | *n* = 2,201 | *n* = 170 | *n* = 480 | *n* = 829 | *n* = 175 | *n* = 264 | *n* = 283 | *n* = 1,787 | *n* = 392 |
| High school or less | 489<br>(22.2%)<sup>H</sup> | 43 (25%) | 124<br>(25.9%)<sup>AG</sup> | 179<br>(21.5%) | 37 (21.4%) | 54<br>(20.6%) | 51 (18.1%) | 354<br>(19.8%) | 131<br>(33.4%)<sup>AH</sup> |
| Trades/College | 663<br>(30.1%)<sup>HG</sup> | 61<br>(35.7%)<sup>G</sup> | 180<br>(37.5%)<sup>ADE</sup><sub>FG</sub> | 240<br>(29%)<sup>G</sup> | 47 (26.8%) | 76<br>(28.7%)<sup>G</sup> | 60 (21.2%) | 500 (28%) | 158<br>(40.1%)<sup>AH</sup> |
| University | 1,046<br>(47.5%)<sup>IBC</sup> | 67 (39.2%) | 174<br>(36.3%) | 409<br>(49.3%)<sup>B</sup> | 91 (51.8%)<sup>BC</sup> | 134<br>(50.6%)<sup>BC</sup> | 172<br>(60.6%)<sup>ABC</sup><br>DF | 932<br>(52.1%)<sup>AI</sup> | 103<br>(26.2%) |
| **Employment** | *n* = 2,201 | *n* = 170 | *n* = 480 | *n* = 829 | *n* = 175 | *n* = 264 | *n* = 283 | *n* = 1,787 | *n* = 392 |
| Employed (Full-time, Part-time, Self-employed) | 1,642<br>(74.6%)<sup>I</sup> | 120<br>(70.7%) | 368<br>(76.8%) | 601<br>(72.5%) | 128 (73.2%) | 197<br>(74.8%) | 227<br>(80.1%)<sup>ABD</sup> | 1,371<br>(76.7%)<sup>AI</sup> | 257<br>(65.6%) |
| *Working full-time (30 or more hours per week)* | 1299<br>(59%)<sup>ID</sup> | 104<br>(61.2%) | 305<br>(63.5%)<sup>AD</sup> | 452<br>(54.5%) | 104 (59.5%) | 161<br>(61.2%) | 172<br>(60.9%) | 1,097<br>(61.4%)<sup>AI</sup> | 192<br>(48.9%) |
| *Working part-time (less than 30 hours per week)* | 205<br>(9.3%)<sup>BC</sup> | 9 (5.1%) | 35 (7.2%) | 100<br>(12.1%)<sup>ABC</sup><br>EF | 12 (6.7%) | 19<br>(7.1%) | 31<br>(10.9%)<sup>B</sup> | 170 (9.5%) | 32<br>(8.2%) |
| *Self-employed* | 138<br>(6.3%) | 7 (4.3%) | 29 (6%) | 49 (6%) | 12 (6.9%) | 17<br>(6.4%) | 23 (8.3%) | 104 (5.8%) | 34<br>(8.5%) |
| Unemployed (looking for work) | 161<br>(7.3%)<sup>C</sup> | 12 (7.2%) | 24 (5%) | 76<br>(9.2%)<sup>ACF</sup> | 14 (8%) | 15<br>(5.7%) | 19 (6.9%) | 129 (7.2%) | 31 (8%) |
| Not employed/not in workforce | 348<br>(15.8%)<sup>HG</sup> | 33<br>(19.2%)<sup>G</sup> | 73 (15.2%) | 132<br>(15.9%) | 31 (17.4%) | 47<br>(17.8%)<sup>G</sup> | 33 (11.6%) | 253<br>(14.2%) | 89<br>(22.6%)<br>AH |
| Other | 31<br>(1.4%)<sup>H</sup> | 5 (2.9%) | 13<br>(2.6%)<sup>AF</sup> | 10 (1.2%) | - | - | - | 17 (0.9%) | 14<br>(3.6%)<sup>A</sup><br>H |
| **Household income\*** | *n* = 2,201 | *n* = 170 | *n* = 480 | *n* = 829 | *n* = 175 | *n* = 264 | *n* = 283 | *n* = 1,787 | *n* = 392 |
| Under \$20,000 | 169<br>(7.7%)<sup>HCG</sup> | 18<br>(10.5%)<sup>CG</sup> | 26 (5.4%) | 79<br>(9.5%)<sup>ACG</sup> | 18 (10.1%)<sup>CG</sup> | 18<br>(6.7%) | 11 (3.8%) | 122 (6.8%) | 44<br>(11.1%)<br>AH |
| \$20,000 to \$29,999 | 200<br>(9.1%)<sup>HE</sup> | 15 (8.8%) | 51<br>(10.6%)<sup>E</sup> | 75 (9.1%)<sup>E</sup> | 9 (5%) | 23<br>(8.6%) | 27 (9.6%)<sup>E</sup> | 150 (8.4%) | 46<br>(11.7%) |
| \$30,000 to \$39,999 | 212<br>(9.6%) | 16 (9.6%) | 53 (11%) | 76 (9.2%) | 17 (9.5%) | 22<br>(8.5%) | 27 (9.6%) | 175 (9.8%) | 36<br>(9.2%) |
| \$40,000 to \$49,999 | 278<br>(12.6%) | 21 (12.2%) | 63 (13%) | 116<br>(13.9%) | 21 (12.2%) | 27<br>(10.4%) | 30 (10.6%) | 225<br>(12.6%) | 52<br>(13.2%) |
| \$50,000 to \$59,999 | 356<br>(16.2%) | 27 (16%) | 83 (17.2%) | 133 (16%) | 23 (13.4%) | 43<br>(16.3%) | 47 (16.5%) | 290<br>(16.2%) | 64<br>(16.3%) |
| \$60,000 to \$69,999 | 396<br>(18%)<sup>ID</sup> | 31 (18.4%) | 89 (18.6%) | 125<br>(15.1%) | 38 (21.5%)<sup>D</sup> | 50<br>(18.8%) | 63<br>(22.3%)<sup>D</sup> | 335<br>(18.8%)<sup>AI</sup> | 55<br>(14%) |
| \$70,000 to \$79,999 | 349<br>(15.9%) | 25 (14.9%) | 69 (14.4%) | 132<br>(15.9%) | 32 (18.4%) | 40<br>(15.3%) | 50 (17.8%) | 289<br>(16.2%) | 58<br>(14.8%) |
| \$80,000 to \$89,999 | 242 (11%) | 16 (9.7%) | 46 (9.6%) | 93<br>(11.2%) | 18 (10.1%) | 41<br>(15.4%)<sup>A</sup><br>CG | 28 (9.9%) | 202<br>(11.3%) | 38<br>(9.6%) |
| **Marital status** | *n* = 2,201 | *n* = 170 | *n* = 480 | *n* = 829 | *n* = 175 | *n* = 264 | *n* = 283 | *n* = 1,787 | *n* = 392 |
| Never married | 304<br>(13.8%)<sup>G</sup> | 34<br>(19.8%)<sup>AEF</sup><br>G | 84<br>(17.4%)<sup>AEF</sup><br>G | 113<br>(13.7%) | 17 (9.9%) | 29 (11%) | 27 (9.6%) | 248<br>(13.9%) | 55<br>(14%) |
| Married/Common law | 1,712<br>(77.8%)<sup>C</sup> | 127<br>(74.4%) | 351<br>(73.2%) | 643<br>(77.5%) | 145<br>(83.1%)<sup>ABC</sup> | 216<br>(81.9%)<sup>C</sup> | 230<br>(81.1%)<sup>C</sup> | 1,404<br>(78.5%) | 291<br>(74%) |
| Separated/Divorced/Widowed | 175 (8%)<sup>H</sup> | 10 (5.8%) | 41 (8.6%) | 71 (8.5%) | 11 (6.5%) | 18<br>(6.7%) | 24 (8.6%) | 127 (7.1%) | 45<br>(11.5%)<br>AH |
| Other | - | - | - | - | - | - | - | - | - |
| Prefer not to answer | 7 (0.3%) | - | - | - | - | - | - | 5 (0.3%) | - |
| **Age of children** | *n* = 2,201 | *n* = 170 | *n* = 480 | *n* = 829 | *n* = 175 | *n* = 264 | *n* = 283 | *n* = 1,787 | *n* = 392 |
| Under age 12 | 2,201<br>(100%) | 170<br>(100%) | 480<br>(100%) | 829<br>(100%) | 175 (100%) | 264<br>(100%) | 283<br>(100%) | 1,787<br>(100%) | 392<br>(100%) |
| 12-17 years of age | 399<br>(18.1%)<sup>BG</sup> | 19 (11.4%) | 105<br>(21.8%)<sup>ABE</sup><br>G | 162<br>(19.5%)<sup>BG</sup> | 27 (15.3%) | 50<br>(18.9%)<sup>B</sup> | 37 (13%) | 312<br>(17.5%) | 82<br>(21%) |
| 18 years of age or older | 90<br>(4.1%)<sup>H</sup> | 9 (5.5%) | 25 (5.2%)<sup>F</sup> | 32 (3.9%) | 7 (3.8%) | 7 (2.5%) | 10 (3.4%) | 61 (3.4%) | 27<br>(7%)<sup>AH</sup> |
| **Private dental insurance\*\*** | *n* = 2,201 | *n* = 170 | *n* = 480 | *n* = 829 | *n* = 175 | *n* = 264 | *n* = 283 | *n* = 1,787 | *n* = 392 |
| Yes | 708<br>(32.2%)<sup>HG</sup> | 73<br>(42.7%)<sup>ACD</sup><br>FG | 152<br>(31.7%)<sup>G</sup> | 272<br>(32.8%)<sup>G</sup> | 63 (35.8%)<sup>G</sup> | 79<br>(29.8%) | 70 (24.7%) | 549<br>(30.7%) | 143<br>(36.5%)<br>AH |
| No | 1,493<br>(67.8%)<sup>IB</sup> | 97 (57.3%) | 328<br>(68.3%)<sup>B</sup> | 557<br>(67.2%)<sup>B</sup> | 112 (64.2%) | 185<br>(70.2%)<sup>B</sup> | 213<br>(75.3%)<sup>ABC</sup><br>DE | 1,238<br>(69.3%)<sup>AI</sup> | 249<br>(63.5%) |
| **Ethnicity(ies)** | *n* = 2,201 | *n* = 170 | *n* = 480 | *n* = 829 | *n* = 175 | *n* = 264 | *n* = 283 | *n* = 1,787 | *n* = 392 |
| Western European (UK, Spain, Portugal, France, Italy, Germany, Austria, Switzerland, etc.) | 567<br>(25.8%)<sup>H</sup> | 39 (22.9%) | 128<br>(26.6%) | 202<br>(24.3%) | 50 (28.4%) | 63<br>(23.8%) | 86 (30.4%) | 440<br>(24.6%) | 126<br>(32.1%)<sup>AH</sup> |
| Eastern European (Poland, Hungary, Romania, Ukraine, Russia, etc.) | 153<br>(7%)<sup>CD</sup> | 13 (7.6%)<sup>C</sup> | 9 (1.8%) | 46 (5.6%)<sup>C</sup> | 23<br>(13.4%)<sup>ACD</sup> | 32<br>(12.1%)<sup>A</sup><br>CD | 30<br>(10.6%)<sup>ACD</sup> | 123 (6.9%) | 30<br>(7.6%) |
| Canadian Indigenous (First Nations, Métis, Inuit (Inuk), etc.) | 291<br>(13.2%)<sup>C</sup> | 31<br>(18.3%)<sup>C</sup> | 43 (9%) | 106<br>(12.8%)<sup>C</sup> | 29 (16.4%)<sup>C</sup> | 40<br>(15.2%)<sup>C</sup> | 42<br>(14.7%)<sup>C</sup> | 236<br>(13.2%) | 54<br>(13.8%) |
| NET ETHNIC (African, Middle Eastern, South Asian, Southeast Asian, East Asian, South/Central/Latin American, and West Indian)<sup>§</sup> | 786<br>(35.7%)<sup>IBC</sup> | 35 (20.6%) | 92 (19.3%) | 372<br>(44.8%)<sup>ABC</sup><br>F | 70 (39.8%)<sup>BC</sup> | 99<br>(37.5%)<sup>BC</sup> | 118<br>(41.5%)<sup>ABC</sup> | 717<br>(40.1%)<sup>AI</sup> | 47<br>(12%) |
| Other<sup>§§</sup> | 275<br>(12.5%)<sup>HD</sup><br>EG | 33<br>(19.6%)<sup>ADE</sup><br>FG | 117<br>(24.5%)<sup>ADE</sup><br>FG | 79 (9.6%)<sup>G</sup> | 10 (5.8%) | 28<br>(10.7%)<sup>E</sup><br>G | 7 (2.4%) | 189<br>(10.6%) | 86<br>(21.9%)<sup>AH</sup> |
| Don't know | 127<br>(5.8%)<sup>HDEG</sup> | 14 (8%)<sup>EG</sup> | 54<br>(11.2%)<sup>ADE</sup><br>FG | 37 (4.5%) | - | 10<br>(3.9%) | 9 (3.1%) | 84 (4.7%) | 43<br>(11%)<sup>AH</sup> |
| Prefer not to answer | 90<br>(4.1%)<sup>HDEF</sup> | 9 (5.6%)<sup>F</sup> | 42<br>(8.8%)<sup>ADEFG</sup> | 22 (2.6%) | - | - | 9 (3.1%) | 62 (3.5%) | 27<br>(6.9%)<sup>A</sup><br>H |

Note: Comparison Groups: ABCDEFG/AH!; Paired/overlap T-test for means, paired/overlap Z-test for percentages. Uppercase letters indicate significance at $p \leq 0.05$ .
\* Last year's total household income, before taxes, for the respondent and their spouse/common-law partner; not including any income received via universal childcare benefit.
\*\* Private insurance for the respondent and their family, either through an employer, pension plan, or individual benefits plan; not including any coverage received from any provincial or federal dental plans.
§ NET ETHNIC: detailed information can be provided upon request.
§§ Other includes Canadian (general), White/Caucasian, Mixed (general), Quebecois, North American, and None.

A telephone recruit-to-online approach was used to reach those residing in remote communities who met the eligibility criteria. A weighting scheme was applied aiming to align the sample size with the distribution of the Canadian population by region, based on the Census 2021 from Statistics Canada. Prior to the launch of the survey, on March 2, 2023, the Strategic Counsel conducted a pre-test with 32 respondents (11 in French and 21 in English) (Health Canada 2023).

The survey was administered from March 3^rd^ until March 30^th^, 2023, and the detailed methodology used for sample design can be found in the Canada Dental Benefit Baseline Survey Final Report (HC POR —22-32).(Health Canada 2023) Data from this study were accessed from the Government of Canada’s “Library and Archives Canada collection” https://epe.bac-lac.gc.ca/100/200/301/pwgsc-tpsgc/por-ef/health/2023/126-22-e/index.html.

Institutional ethics approval was not required as this study utilized aggregate and anonymized data that did not contain personal identifiers. Specific variables that were evaluated in this study included demographic characteristics of the respondents (e.g., age, gender, education, and employment), attitudes towards dental care, access to and use of dental services, barriers to accessing dental care for children, and awareness and views on the Interim CDB. The analysis were done by province/region and by place of residence (urban: a city or large town or rural: outside a city or a large town). The place of residence was determined based on the postal code or it was self-reported by those who preferred not to provide a postal code. Values less than five were suppressed for confidentiality purposes. Paired/overlap t-test for means and paired/overlap z-test for percentages were performed, with statistical significance at *p* ≤ 0.05.

## Results

### Characteristics of the participants

The majority of participants were between 25-44 years old (78%), female (62.1%), married or in common-law relationships (77.8%), employed (74.6%), lacked private dental insurance (67.8%), had university (47.5%) or trades/college (30.1%) education level, lived in urban centres (81.2%), and were from Ontario (37.7%) (Table 1). There was a wide distribution of household income levels, with nearly 45% earning $60,000 or more. The distribution of income levels showed some regional variation, with the Atlantic region having a higher proportion of participants earning under $40,000. The ethnic backgrounds of participants was diverse, with the largest group being Western European (25.8%), while another 13.2% identified as Canadian Indigenous.

### View and concern about affordability of and access to dental care

The majority of the participants (90.9%) were very or somewhat concerned about the affordability of dental care in Canada, with regional responses ranging from 88% in Atlantic Canada to 93.3% in Manitoba and Saskatchewan. There was no difference in responses between urban and rural dwellers. Further, 80.9% were very or somewhat concerned about accessing the services of a dentist or oral health care professional (Table 2), with regional responses ranging from 73.9% in Atlantic Canada to 83.9% in Manitoba and Saskatchewan. Interestingly, those living in urban areas were more likely to report concerns about accessing services of a dentist or oral health care professional than those from rural areas (82.3% vs. 74.2%).

**Table 2.** Level of concern about affordability of dental care in Canada and accessing the services of a dentist or oral health care professional (e.g., dental assistant, dental hygienist, dental surgeon, etc.), by region.

|  | Total<br>(A) | Atlantic<br>(B) | Quebec<br>(C) | Ontario<br>(D) | Manitoba/<br>Saskatchewan<br>(E) | Alberta<br>(F) | British<br>Columbia/<br>North<br>(G) | URBAN<br>(H) | RURAL<br>(I) |
| --- | --- | --- | --- | --- | --- | --- | --- | --- | --- |
| **Affordability of dental care** | *n* = 2,201 | *n* = 170 | *n* = 480 | *n* = 829 | *n* = 175 | *n* = 264 | *n* = 283 | *n* = 1,787 | *n* = 392 |
| VERY/SOMEWHAT CONCERNED | 2,000<br>(90.9%)<sup>C</sup> | 157 (92.3%) | 422 (88%) | 759<br>(91.6%)<sup>C</sup> | 163 (93.3%)<sup>C</sup> | 235<br>(88.9%) | 263<br>(93.1%)<sup>C</sup> | 1,623<br>(90.8%) | 357 (91%) |
| *Very concerned* | 1,339<br>(60.8%) | 99 (58.4%) | 283<br>(58.9%) | 512<br>(61.8%) | 96 (55%) | 169<br>(64%)<sup>E</sup> | 179<br>(63.3%) | 1,090<br>(61%) | 234 (59.6%) |
| *Somewhat concerned* | 661 (30%)<sup>F</sup> | 58 (33.9%)<sup>F</sup> | 139<br>(29.1%) | 247<br>(29.8%) | 67 (38.2%)<sup>ACDF</sup> | 66<br>(24.8%) | 84 (29.8%) | 533<br>(29.8%) | 123 (31.4%) |
| NOT THAT/NOT AT ALL CONCERNED | 201 (9.1%) | 13 (7.7%) | 58<br>(12%)<sup>ADEG</sup> | 70 (8.4%) | 12 (6.7%) | 29<br>(11.1%) | 20 (6.9%) | 165<br>(9.2%) | 35 (9%) |
| *Not that concerned* | 165 (7.5%) | 13 (7.4%) | 49<br>(10.2%)<sup>AEG</sup> | 59 (7.1%) | 9 (4.9%) | 21 (7.9%) | 15 (5.2%) | 131<br>(7.3%) | 33 (8.4%) |
| *Not concerned at all* | 36 (1.6%)<sup>IB</sup> | - | 9 (1.8%)<sup>B</sup> | 11 (1.3%) | - | 9 (3.2%)<sup>B</sup> | 5 (1.7%) | 34 (1.9%)<sup>I</sup> | - |
| **Accessing the services of a dentist or oral health care professional** | *n* = 2,201 | *n* = 170 | *n* = 480 | *n* = 829 | *n* = 175 | *n* = 264 | *n* = 283 | *n* = 1,787 | *n* = 392 |
| VERY/SOMEWHAT CONCERNED | 1,781<br>(80.9%)<sup>IB</sup> | 126 (73.9%) | 378<br>(78.7%) | 679<br>(81.9%)<sup>B</sup> | 147 (83.9%)<sup>B</sup> | 218<br>(82.6%)<sup>B</sup> | 234<br>(82.5%)<sup>B</sup> | 1,470<br>(82.3%)<sup>AI</sup> | 291 (74.2%) |
| *Very concerned* | 1,001<br>(45.5%) | 68 (40%) | 200<br>(41.7%) | 388<br>(46.8%) | 87 (49.7%) | 121<br>(45.9%) | 137<br>(48.3%) | 823<br>(46.1%) | 162 (41.3%) |
| *Somewhat concerned* | 780 (35.4%) | 58 (33.9%) | 178<br>(37.1%) | 291<br>(35.1%) | 60 (34.1%) | 97<br>(36.7%) | 97 (34.3%) | 647<br>(36.2%) | 129 (32.9%) |
| NOT THAT/NOT AT ALL CONCERNED | 420<br>(19.1%)<sup>H</sup> | 44<br>(26.1%)<sup>ADEFG</sup> | 102<br>(21.3%) | 150<br>(18.1%) | 28 (16.1%) | 46<br>(17.4%) | 49 (17.5%) | 317<br>(17.7%) | 101<br>(25.8%)<sup>AH</sup> |
| *Not that concerned* | 334<br>(15.2%)<sup>H</sup> | 34 (19.7%)<sup>E</sup> | 88<br>(18.3%)<sup>AE</sup> | 117<br>(14.1%) | 21 (12.1%) | 38<br>(14.2%) | 38 (13.4%) | 248<br>(13.9%) | 86<br>(21.9%)<sup>AH</sup> |
| *Not concerned at all* | 86 (3.9%) | 11 (6.4%) | 14 (3%) | 34 (4%) | 7 (4%) | 8 (3.2%) | 12 (4.1%) | 70 (3.9%) | 15 (3.9%) |

Note: Comparison Groups: ABCDEFG/AHI; Paired/overlap T-test for means, paired/overlap Z-test for percentages. Uppercase letters indicate significance at $p \leq 0.05$ .

Almost all participants (97.2%) indicated that regular dental visits for children is important, with similar responses across regions and between urban and rural dwellers (Table 3). When asked, a majority of participants reported they would schedule more regular dental care appointments for their children (79.9%) or themselves (82.1%) if they had a bit of extra money. However, there were some regional variations for scheduling more regular dental care appointments for children (ranging from 69.9% in Atlantic Canada to 85.6% in Manitoba and Saskatchewan) and parents (ranging from 73.4% in Atlantic Canada to 83.5% in Manitoba and Saskatchewan). Overall, 58.7% or respondents indicated that they only schedule dental care appointments for their children when absolutely necessary, with regional variations (ranging from 49.5% in Atlantic Canada to 72.0% in Manitoba and Saskatchewan), and urban and rural differences (60.2% vs. 51.1%). Additionally, 94.7% of the participants strongly or somewhat agreed that, ideally, they would like their children and themselves to receive regular dental care (Table 3).

**Table 3.**
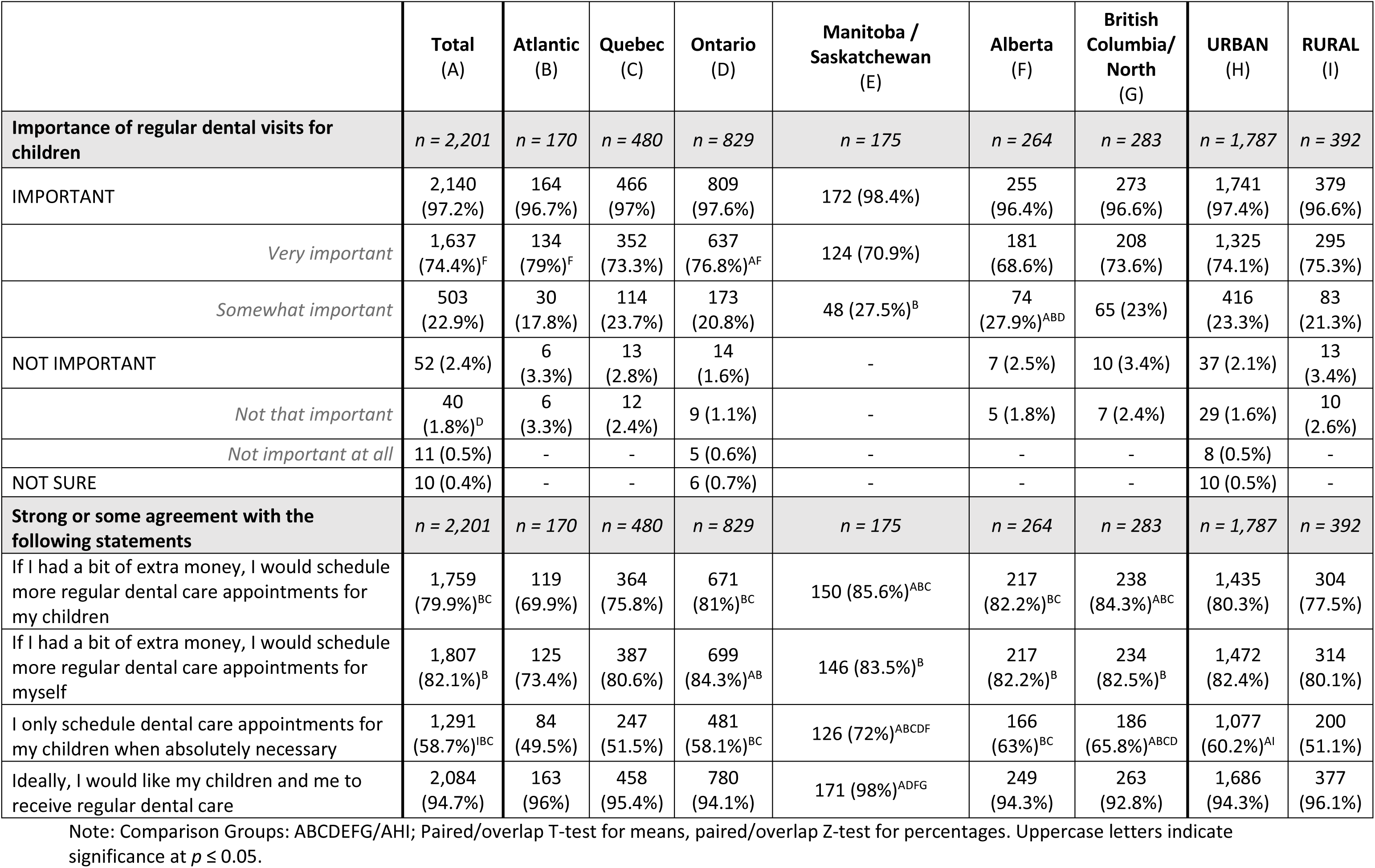
Views on dental care for children.

### Access to dental services for children

Overall, 51.3% of the participants reported having access to a dentist or oral health professional for their whole family and 18.5% having access only for their children (Table 4). The Atlantic region had the highest proportion of respondents (59.9%) reporting they had access to dental services for the whole family, while Manitoba and Saskatchewan had the lowest proportion of residents reporting access to dental services for the entire family (47.3%). Surprisingly, more participants in rural regions reported having access to dental services for the whole family than those from urban areas (58.1% vs. 49.7%).

**Table 4.**
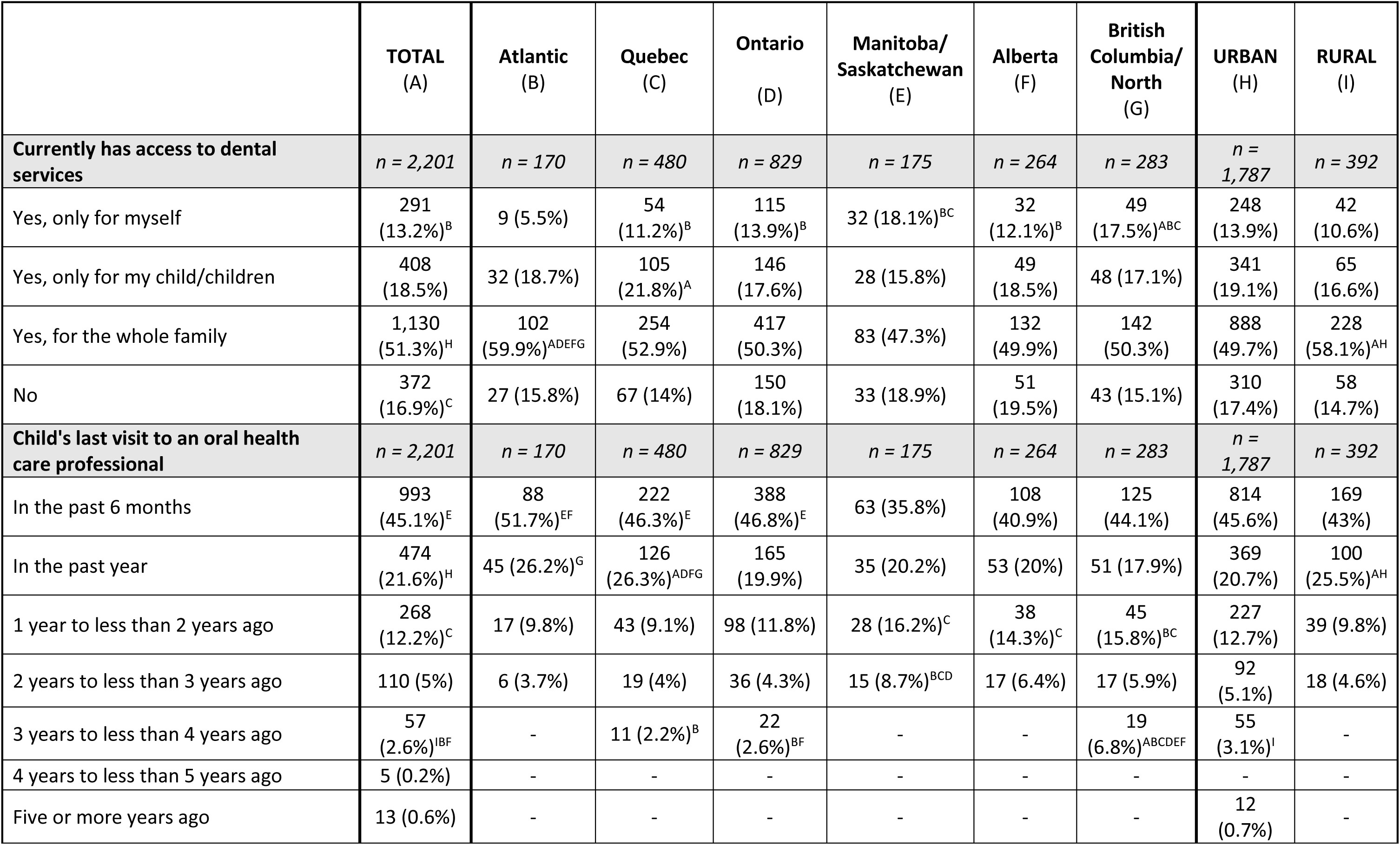
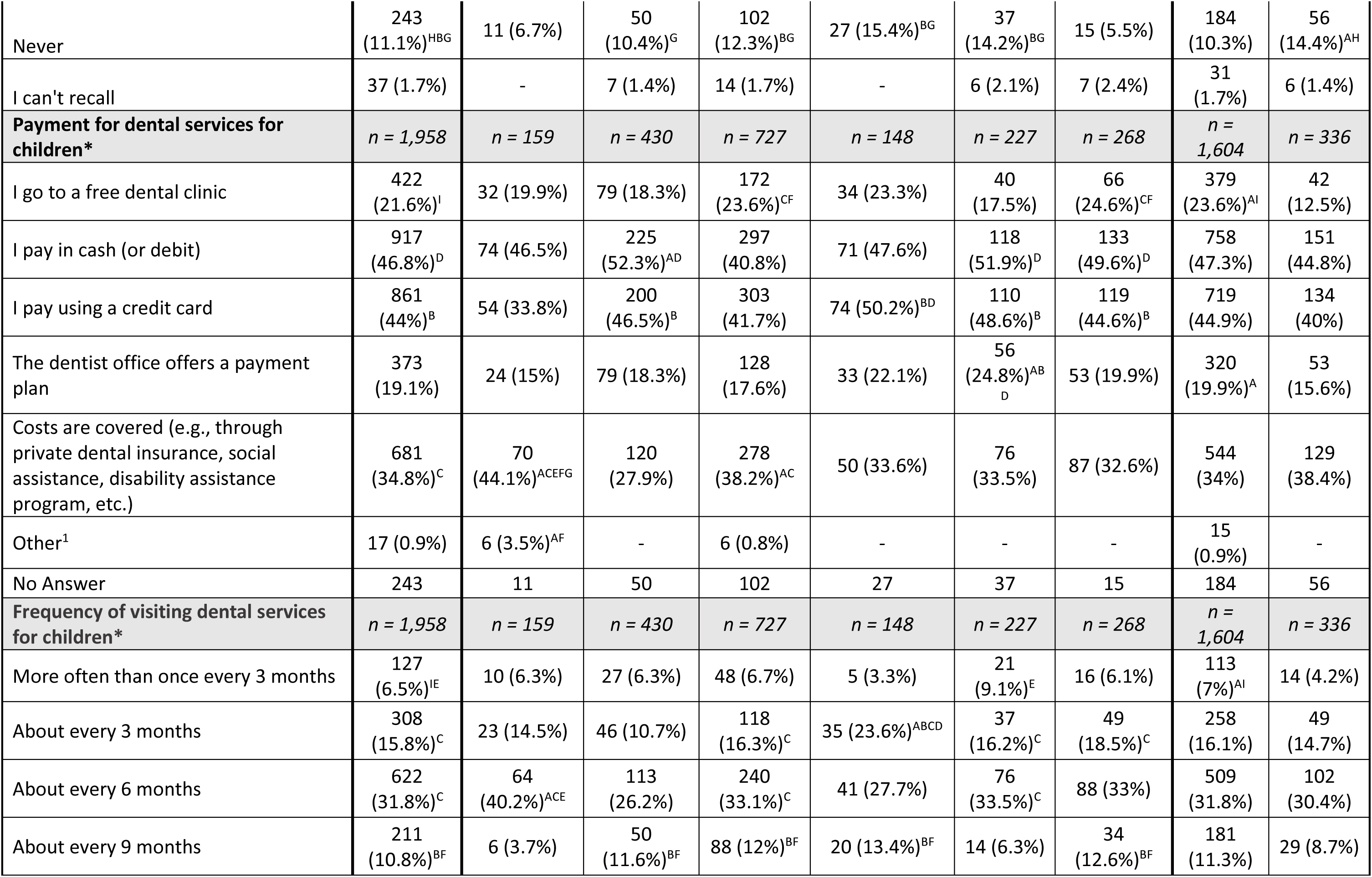
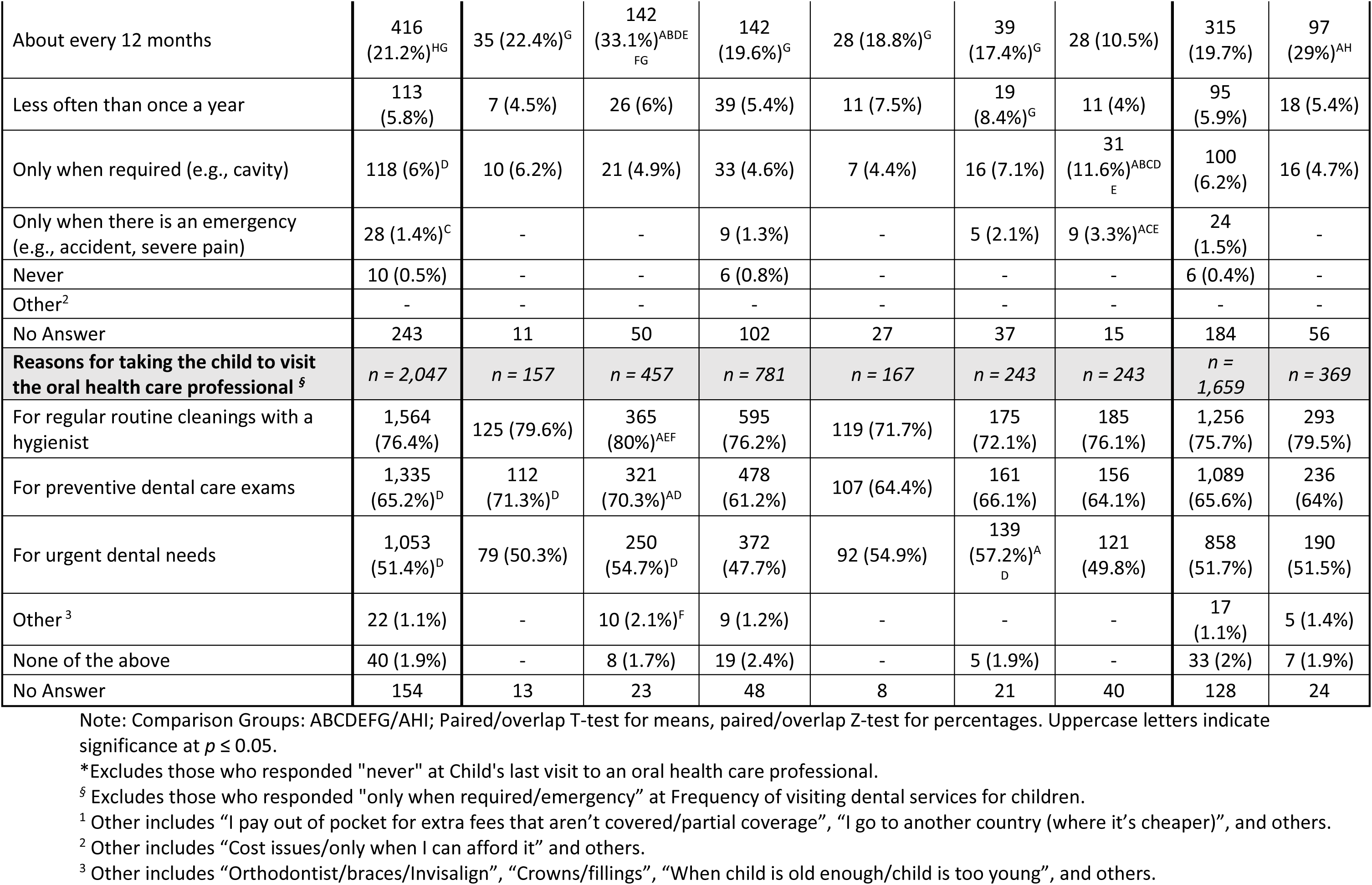
Access to and use of dental services for children oral health care, by region.

Furthermore, 45.1% of participants reported taking their child to a dental office or oral health professional within the past 6 months prior to the survey (Table 4), with results ranging from 51.7% in Atlantic Canada to 35.8% in Manitoba and Saskatchewan. There was no significant difference between urban and rural respondents. Overall, 66.7% reported that their child last visited an oral health care professional within the last year or less, with results ranging from 77.9% in Atlantic Canada to 56.0% in Manitoba and Saskatchewan. More parents from rural regions (25.5%) reported their child last visited an oral health care provided within the past year compared to urban regions (20.7%). However, 11.1% of the parents reported that they had never taken their child/children to an oral health care professional. This proportion was highest in Manitoba/Saskatchewan (15.4%), followed by Alberta (14.2%), Ontario (12.3%), and in rural regions (14.4%). Only 34.8% of respondents reported having the costs of dental services for children covered by an insurance or government program, with many having to pay for these services out of pocket. More than half of the parents from Quebec (52.3%) and Alberta (51.9%) reported paying in cash or debit for their child’s dental services. British Columbia (24.6%), Manitoba and Saskatchewan (23.6%), and urban dwellers (23.6%) had the highest proportion of parents taking their children to free dental clinics.

Overall, 31.8% of the respondents took their children to see a dentist or other oral health care professional every 6 months, and the most common reasons for taking their child to visit an oral health care professional were regular routine cleanings, preventive dental exams and urgent dental needs (Table 4).When asked about the barriers preventing parents from taking their child to see an oral health care professional on a regular basis, participants selected the cost of the service (41%), lack of insurance (28%), cost of transportation (25%), cannot miss school or work (19%), anxiety or fear (17%), among others (Figure 1).

**Figure 1.**
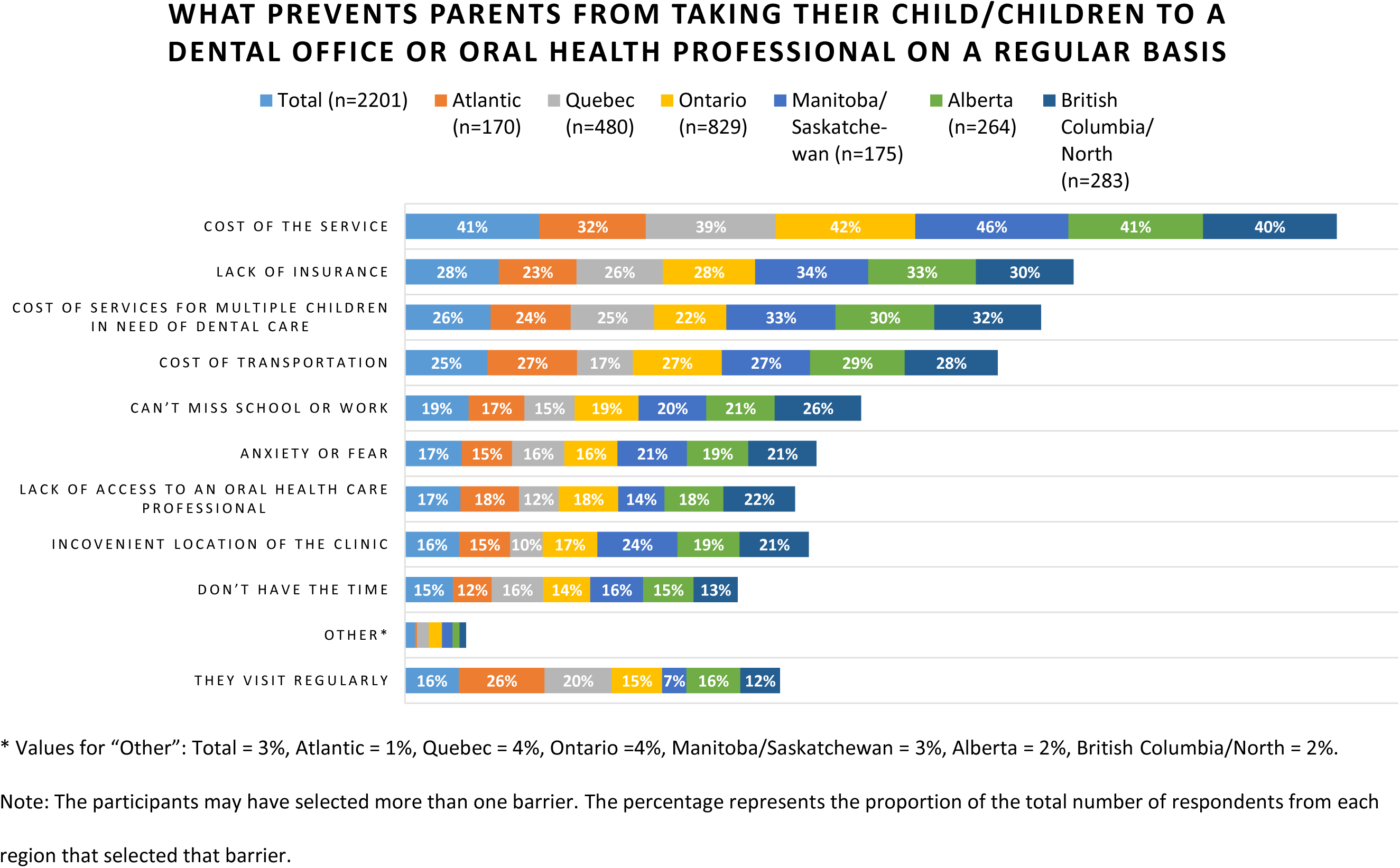
Barriers to accessing dental care for children.

### Awareness of and support for the Interim CDB

Six in ten parents reported hearing about the Interim CDB, but only 22.4% were well informed about the program. Manitoba/Saskatchewan (37.8%) and Atlantic Canada (37.5%) had the highest proportion of people unaware about the Interim CDB. Interestingly, there were higher proportion of parents aware about the program in rural areas (67.9%) compared to urban areas (63.3%). At the same time, parents from rural areas were less likely to apply for the CDB than parents from urban areas. More than 70% of the respondents from British Columbia/North had applied or were planning to apply for the program. Atlantic Canada had the lowest proportion of participants (54.5%) who had applied or were planning to apply for the program (Table 5). Over 60% of the participants without access to dental insurance had heard about the program, and more than 70% were planning to apply or had applied already (Table 5). The majority of parents (87%) showed support for the interim CDB program with the greatest support for the interim CDB coming from Manitoba and Saskatchewan (90.4%).

**Table 5.** Awareness about the Interim Canada Dental Benefit and number of applicants.

|  | Total<br>(A) | Atlantic<br>(B) | Quebec<br>(C) | Ontario<br>(D) | Manitoba /<br>Saskatchewan<br>(E) | Alberta<br>(F) | British<br>Columbia/<br>North<br>(G) | URBAN<br>(H) | RURAL<br>(I) | Access to dental<br>Insurance |  |
| --- | --- | --- | --- | --- | --- | --- | --- | --- | --- | --- | --- |
|  |  |  |  |  |  |  |  |  |  | YES<br>(J) | NO<br>(K) |
| **Heard about the Interim**<br>**CDB?** | *n* = 2,201 | *n* = 170 | *n* = 480 | *n* = 829 | *n* = 175 | *n* = 264 | *n* = 283 | *n* = 1,787 | *n* = 392 | *n* = 708 | *n* = 1,493 |
| NO | 787<br>(35.7%)<sup>C</sup> | 64<br>(37.5%) | 151<br>(31.5%) | 301<br>(36.4%) | 66 (37.8%) | 99<br>(37.4%) | 106<br>(37.4%) | 656<br>(36.7%)<sup>A</sup> | 126<br>(32.1%) | 198<br>(28%) | 588<br>(39.4%)<sup>AJ</sup> |
| YES | 1,414<br>(64.3%)<sup>H</sup> | 106<br>(62.5%) | 329<br>(68.5%)<sup>A</sup> | 528<br>(63.6%) | 109 (62.2%) | 165<br>(62.6%) | 177<br>(62.6%) | 1,132<br>(63.3%) | 266<br>(67.9%) | 510<br>(72%)<sup>AK</sup> | 905<br>(60.6%) |
| *Yes, I have heard of it,*<br>*but don't know much about*<br>*the program* | 922<br>(41.9%)<sup>HG</sup> | 72<br>(42.6%) | 213<br>(44.3%)<sup>G</sup> | 347<br>(41.8%) | 80 (45.5%)<sup>G</sup> | 109<br>(41.2%) | 102 (36%) | 724<br>(40.5%) | 186<br>(47.4%)<sup>AH</sup> | 378<br>(53.4%)<sup>AK</sup> | 544<br>(36.5%) |
| *Yes, I have heard of it*<br>*and am well informed*<br>*about the program* | 492<br>(22.4%)<sup>E</sup> | 34<br>(19.9%) | 116<br>(24.2%)<sup>E</sup> | 181<br>(21.8%) | 29 (16.6%) | 56<br>(21.3%) | 76 (26.7%)<sup>E</sup> | 408<br>(22.8%) | 80<br>(20.5%) | 132<br>(18.6%) | 360<br>(24.1%)<sup>AJ</sup> |
| **Applied or planning to**<br>**apply for the Interim CDB?** | *n* = 2,201 | *n* = 170 | *n* = 480 | *n* = 829 | *n* = 175 | *n* = 264 | *n* = 283 | *n* = 1,787 | *n* = 392 | *n* = 708 | *n* = 1,493 |
| NO, I have not applied and<br>do not plan to | 487<br>(22.1%)<sup>D</sup> | 48<br>(28.5%)<sup>ADG</sup> | 116<br>(24.2%)<sup>D</sup> | 159<br>(19.2%) | 45 (25.6%) | 63<br>(23.8%) | 55 (19.6%) | 384<br>(21.5%) | 97<br>(24.7%) | 287<br>(40.5%)<sup>AK</sup> | 200<br>(13.4%) |
| Not sure | 246<br>(11.2%)<sup>HG</sup> | 29<br>(17%)<sup>ADG</sup> | 55<br>(11.4%)<sup>G</sup> | 94<br>(11.3%)<sup>G</sup> | 19 (10.8%) | 29<br>(11%) | 20 (7.2%) | 178<br>(10%) | 66<br>(16.8%)<sup>AH</sup> | 117<br>(16.5%)<sup>AK</sup> | 129<br>(8.6%) |
| YES | 1,468<br>(66.7%)<sup>IB</sup> | 93<br>(54.5%) | 309<br>(64.3%)<sup>B</sup> | 576<br>(69.5%)<sup>AB</sup> | 111 (63.6%) | 172<br>(65.2%)<sup>B</sup> | 207<br>(73.3%)<sup>ABCEF</sup> | 1,225<br>(68.5%)<sup>AI</sup> | 230<br>(58.5%) | 304<br>(43%) | 1,164<br>(77.9%)<sup>AJ</sup> |
| *Yes, I have applied* | 400<br>(18.2%)<sup>IBF</sup> | 21<br>(12.3%) | 92<br>(19.2%)<sup>BF</sup> | 150<br>(18.1%)<sup>B</sup> | 32 (18.3%) | 36<br>(13.5%) | 70<br>(24.6%)<sup>ABDF</sup> | 345<br>(19.3%)<sup>AI</sup> | 52<br>(13.4%) | 59<br>(8.3%) | 342<br>(22.9%)<sup>AJ</sup> |
| *Yes, I am planning to*<br>*apply* | 1,068<br>(48.5%) | 72<br>(42.2%) | 216<br>(45.1%) | 426<br>(51.4%)<sup>ABC</sup> | 79 (45.3%) | 136<br>(51.7%)<sup>B</sup> | 138<br>(48.7%) | 880<br>(49.2%) | 177<br>(45.2%) | 246<br>(34.7%) | 822<br>(55.1%)<sup>AJ</sup> |
| **Support for the Interim**<br>**CDB** | *n* = 2,201 | *n* = 170 | *n* = 480 | *n* = 829 | *n* = 175 | *n* = 264 | *n* = 283 | *n* = 1,787 | *n* = 392 | *n* = 708 | *n* = 1,493 |
| SUPPORT | 1,915<br>(87%) | 145<br>(85.2%) | 416<br>(86.8%) | 726<br>(87.6%) | 158 (90.4%)<sup>F</sup> | 223<br>(84.4%) | 247<br>(87.3%) | 1556<br>(87.1%) | 340<br>(86.6%) | 595<br>(84%) | 1,321<br>(88.5%)<sup>AJ</sup> |
| *Strongly support* | 1,404<br>(63.8%)<sup>F</sup> | 119<br>(70%)<sup>F</sup> | 311<br>(64.7%)<sup>F</sup> | 541<br>(65.2%)<sup>F</sup> | 107 (61.1%) | 151<br>(57.3%) | 175<br>(61.9%) | 1,136<br>(63.5%) | 253<br>(64.4%) | 425<br>(60.1%) | 979<br>(65.5%)<sup>AJ</sup> |
| *Somewhat support* | 512<br>(23.2%)<sup>B</sup> | 26<br>(15.2%) | 106<br>(22%)<sup>B</sup> | 185<br>(22.4%)<sup>B</sup> | 51 (29.3%)<sup>B</sup> | 72<br>(27.1%)<sup>B</sup> | 72 (25.4%)<sup>B</sup> | 421<br>(23.5%) | 87<br>(22.2%) | 169<br>(23.9%) | 342<br>(22.9%) |
| OPPOSE | 59 (2.7%) | - | 13<br>(2.6%) | 18 (2.1%) | 5 (3%) | 9 (3.3%) | 13 (4.5%) | 50 (2.8%) | 9 (2.3%) | 30<br>(4.2%)<sup>AK</sup> | 29 (2%) |
| *Somewhat oppose* | 41 (1.9%) | - | 8 (1.6%) | 12 (1.4%) | - | - | 11 (3.8%)<sup>D</sup> | 36 (2%) | 5 (1.3%) | 22<br>(3.1%)<sup>AK</sup> | 19 (1.3%) |
| *Strongly oppose* | 18 (0.8%) | - | 5 (1%) | 6 (0.7%) | - | 5 (1.8%) | - | 14 (0.8%) | - | 8 (1.1%) | 10 (0.7%) |
| Neither support nor<br>oppose | 196<br>(8.9%)<sup>E</sup> | 19<br>(11.3%)<sup>E</sup> | 44<br>(9.2%) | 76 (9.2%) | 10 (5.6%) | 28<br>(10.6%)<sup>E</sup> | 18 (6.5%) | 156<br>(8.7%) | 38 (9.6%) | 75<br>(10.6%) | 121<br>(8.1%) |
| Don't know | 30 (1.4%) | - | 7 (1.4%) | 9 (1.1%) | - | 5 (1.8%) | 5 (1.7%) | 25 (1.4%) | 5 (1.4%) | 8 (1.2%) | 22 (1.5%) |

Note: Comparison Groups: ABCDEFG/AHI/AJK; Paired/overlap T-test for means, paired/overlap Z-test for percentages. Uppercase letters indicate significance at $p \leq 0.05$ .

## Discussion

This study identified that many Canadian parents report challenges in accessing oral health care for their children under 12 years of age. Specifically, we identified that parents reported concerns about the affordability of dental care, have concerns about accessing services of an oral health professional, only schedule dental appointments for their children when necessary, indicate that they do not have access to an oral health professional, and have to pay for their children’s dental treatment out of pocket. Additionally, parents identified numerous barriers to care, including the cost of services, transportation issues, lack of insurance, absence from work or school, anxiety or fear, inconvenient location of the dental clinic, or lack of access to an oral health care professional. Many of these access-to-care issues appear to be more pronounced in some provinces/regions. Further, respondents clearly expressed tremendous support for the interim CDB policy developed by the federal government as a precursor to the CDCP.

Our analysis revealed substantial concerns regarding the affordability and accessibility of dental care, with over 90% of participants expressing some level of concern. As 32.2% of the participants had private dental insurance, this demonstrates that some people with dental coverage may still experience cost-related barriers, including being underinsured. Additionally, they may be sympathetic to the affordability of oral health care as they may have other family or friends who are uninsured or under-insured and face challenges in accessing oral health care. A recent study analyzing data from the 2017-2018 Canadian Community Health Survey (CCHS) reported that some people living in Ontario, despite having private dental insurance, still face financial barriers to access dental care (Abdelrehim and Singhal 2024). Similarly, data from the 2022 CCHS showed that 16% of Canadian adults with private insurance and 47.4% of those uninsured reported avoidance of dental care because of the cost (Moharrami et al. 2024).

Notably, our analysis underscored a strong consensus on the importance of regular dental visits for children, with 97% of respondents acknowledging its significance. This is consistent with the literature, emphasizing the critical role of early dental care in long-term oral health outcomes (American Academy of Pediatric Dentistry Pediatric Oral Health Research and Policy Center 2014). Studies have shown that regular dental check-ups in childhood can significantly reduce the incidence of oral diseases (Khan et al. 2023), underlining the public health implications of our findings showing that some parents in Canada only take their children to see an oral health care professional when required (e.g., cavity) or when there is an emergency (e.g., pain or infection). This reactive approach to dental health can lead to increased long-term health issues and costs. Programs like the CDB and CDCP could play a pivotal role in shifting public attitudes towards more preventive care approaches, ultimately aiming to reduce the frequency and severity of emergency dental conditions.

Access to oral health care can be influenced by place of residence, with increasing oral health disparities seen in children from rural regions (Schroth et al. 2016). We are unable to explain why rural dwelling participants reported better access to care in this study. However, the findings of our study are not fully generalizable to all rural populations as this study’s sample really did not include participants living in very remote parts of Canada, where access to oral health care is extremely limited, because many of these communities do not have reliable internet service that was needed to complete the online survey. Contrary to common perceptions, as in our study, rural areas sometimes report better access and more frequent dental visits for children than urban areas. This phenomenon may be attributed to targeted rural health initiatives or the lesser competition for dental services in these areas (Sulo et al. 2022). Such findings suggest that rural dental service provision might benefit from unique models of care tailored to their less dense populations.

The Canadian Federal Budget 2024 announced the government’s intent to include more health care professionals working in rural and remote communities (e.g., dentists and dental hygienists) in the Canada Student Loan Forgiveness Program, which is currently only available for doctors and nurses (Government of Canada 2024). Loan repayment programs in the U.S. have been shown to have an important influence on healthcare providers’ choice on where to practice and on their retention in rural communities (Renner et al. 2010). Providing incentives for new graduates to work in under-served communities could help address the current distribution of oral health care providers in Canada and increase access to care in oral health professional shortage areas. Proposals such as mobile dental clinics and enhancements to public transportation could provide more consistent and widespread access to dental care facilities, particularly benefiting those in underserved rural areas (Partido et al. 2021; Patel et al. 2023; Pourat et al. 2020). These measures could alleviate some of the geographical and physical barriers currently hindering consistent dental care access, ensuring that all children and families receive more readily preventative and routine dental services.

Significant disparities in access to dental care were also observed between regions in Canada, with Manitoba and Saskatchewan reporting poorer access and a higher reliance on urgent or emergency dental services when compared to other provinces. In Manitoba and Saskatchewan, there are notable discrepancies in dental care accessibility, highlighted by the higher rates of only seeking care when necessary and a significant portion of children not visiting a dental professional. This reflects broader challenges in access observed in these regions, particularly in rural and remote areas. Despite these challenges, substantial public support for the CDB in these provinces indicates a strong community recognition of the substantial gaps in the existing dental care system, and this is demonstrated by their higher and above national rates of children receiving the Interim CDB (Schroth et al. 2024).

While there is a clear demand for better dental care, organized dentistry in Manitoba has shown resistance to the CDCP. At the beginning of 2024, 44% of the licensed members of the Manitoba Dental Association participated in a survey, and the results showed that the majority (89%) of respondents are unlikely or very unlikely to participate as a provider in the CDCP (Manitoba Dental Association 2024). This shows that while the public reports strong support for public funding of oral health care policy for the uninsured and underinsured, there is a major discrepancy between the public’s support for and dentists’ opposition to the CDCP. Given the existing access issues, this significant opposition suggests a complex interaction between healthcare policy and professional practice norms. This scenario underscores the need for innovative solutions to bridge these gaps.

The introduction of the Interim CDB marked a significant milestone in addressing the affordability and accessibility challenges that have long plagued the Canadian dental care system. However, while our study sheds light on the high levels of concern among parents regarding dental care costs and access, it is imperative to contextualize these findings within the broader spectrum of dental care challenges faced nationally. The CDB and, more recently, the CDCP are designed primarily to mitigate financial barriers to dental care. However, these programs may not adequately address other crucial barriers, such as lack of awareness, accessibility challenges, and cultural appropriateness of services (Hopcraft 2024). These gaps highlight the need for comprehensive strategies that go beyond cost reduction, ensuring inclusive access to dental care across diverse communities. Such an approach enriches the discussion and underscores the transformative potential of these programs in mitigating these long-standing barriers.

The high rate of awareness (64%) and intended application (67%) for the Interim CDB among the participants reflects a proactive stance toward utilizing available resources for dental care. This is indicative of a broader trend observed in recent research, where public health initiatives are increasingly recognized and used by the community, suggesting a positive shift towards more informed health behaviour and resource utilization (Zheng et al. 2024). However, these results also indicate the need for improved communication and outreach efforts to ensure all eligible families are informed and can access these supports as more than one-third appeared unaware of the interim CDB.

### Limitations

This study is not without limitations. As data from an online survey was utilized, it is expected that only people with access to the internet and a computer or mobile device and with basic literacy skills to navigate the internet were surveyed. The online survey methodology might have introduced some selection bias, potentially excluding non-internet users who might face different or additional barriers and those living in more rural and remote regions of the country where internet service may be extremely limited. Furthermore, only people willing to participate in a survey were included, and their attitudes and behaviours may differ from those respondents who refused or were unable or unwilling to complete the survey (Health Canada 2023). The sampling format may have had an impact on the findings in rural and urban areas. Consequently, this cohort may not be representative of the entire target Canadian population. Furthermore, the cross-sectional design limits the ability to ascertain causality between perceived barriers and actual utilization of the CDB and the use of aggregated data did not allow further analyses of associations. The exploration of parental perceptions towards the Interim CDB could greatly benefit from a more diversified methodological framework, possibly incorporating qualitative components to capture a more nuanced understanding of these perceptions.

Continuous community feedback and robust data analysis should guide the evolution of these initiatives moving forward. This approach will ensure that the dental care system in Canada becomes more inclusive and responsive to the needs of its population, paving the way toward eliminating oral health disparities across the country.

## Conclusion

This study has substantiated the presence of significant barriers to dental care access in Canada among parents of children under 12 years of age. The responses identified concerns with affordability of dental care and accessing services of an oral health care professional and lack of access to dental services, with many parents only scheduling dental appointments for their children when absolutely necessary and agreeing that if they had extra money, they would schedule more regular dental appointments for their children. Barriers to accessing oral health care include cost of services and transportation to the dental office, lack of insurance, and others. The widespread support across Canada for the Interim CDB reflects a strong public consensus on the need for more affordable dental care options.

## Data Availability

The data is available at https://epe.bac-lac.gc.ca/100/200/301/pwgsc-tpsgc/por-ef/health/2023/126-22-e/index.html

## Acknowledgements

Funding for this research was made possible from Dr. Schroth’s Canadian Institutes of Health Research Applied Public Health Chair in Public Health Approaches to Improve Access to Oral Health Care and Oral Health Status for Young Children in Canada.

